# Analysis of SARS-CoV-2 Variant-Specific Serum Antibodies Post-Vaccination Utilizing Immortalized Human Hepatocyte-like Cells (HLC) to Assess Development of Protective Immunity

**DOI:** 10.1101/2023.08.08.23293863

**Authors:** Daniel P Collins, Clifford J Steer

## Abstract

**Background:** Our previous studies demonstrated that SARS-CoV-2 spike proteins could bind to hepatocytes via the asialoglycoprotein receptor-1 (ASGR-1) facilitating direct infection by the SARS-CoV-2 virus. Immortalized E12-HLC expressed the phenotypic and biological properties of primary human hepatocytes, including their ability to bind spike proteins via ASGR-1 with exception of the spike 1 protein. This binding could be inhibited by spike protein-specific monoclonal antibodies. We used the same spike-blocking analysis to determine if post-vaccination serum was capable of blocking spike protein binding to HLC. Samples collected from subjects prior to, and post-vaccination were quantified for anti-variant-specific antibody (original wild type, alpha (α), beta (β), gamma (γ) and delta (δ) variants) by a flow cytometry based immunofluorescent assay. Inhibition of variant spike protein binding to HLC and AT-2 (as a known model for spike 1 binding to the ACE-2 receptor) was analyzed by confocal microscopy. This study was designed to investigate the ability of post-vaccination antibodies to mediate immunity to spike S2, and to validate the utility of the E12-HLC in analyzing that immunity.

**Methods:** Serum was collected from 10 individuals pre- and post-vaccination with the J&J, Moderna or Pfizer vaccines. The serum samples were quantified for variant-specific antibodies in a flow cytometry-based immunofluorescent assay utilizing beads coated with biotinylated variant spike proteins (α, β, γ, δ). Presence of variant-specific antibodies was visualized by anti-human IgG-Alexa 488. Inhibition of spike protein binding to cells was analyzed by immunofluorescent confocal analysis. Biotinylated variant spike proteins were preincubated with serum samples and then tested for binding to target cells. Binding was visualized by Streptavidin-Alexa 594. Results were compared to binding of unblocked spike variants.

**Results:** All variant spike proteins tested bound to both the HLC and AT-2 cells. Pre-vaccination serum samples had no detectable reactivity to any of the variant spike proteins and were unable to inhibit binding of the variant spike proteins to either target cell. Post-vaccination serum samples demonstrated a progression of SARS-CoV-2 antibody levels from low early post-vaccination levels to higher levels at 2.5 months after vaccination. Concurrently, serum samples taken at those different timeframes demonstrated that serum obtained from shortly after vaccination were not as effective in blocking spike protein as serum obtained after 2.5 months post-vaccination. Antibody concentrations were not necessarily associated with better blocking of spike protein binding as spike variant-specific serum antibody concentrations varied significantly between subjects and within each subject. It was also demonstrated that vaccination with all the various available vaccines stimulated antibodies that inhibited binding of the available variant spike proteins to both HLC and AT-2 cells.

**Conclusion:** HLC, along with AT-2 cells, provides a useful platform to study the development of protective antibodies that prevent the binding SARS-CoV-2 spike proteins to target cells. It was shown that vaccination with the three available vaccines all elicited serum antibodies that were protective against binding of each of the variant spike proteins to both AT-2 and HLC cells. This study suggests that analysis of immune serum to block spike binding to target cells may be a more useful technique to assess protective immunity than quantitation of gross antibody alone.

## Introduction

SARS-CoV-2 is the virus responsible for the worldwide acute respiratory pandemic that has resulted in over 1 million COVID-19 deaths in the United States alone and is still contributing to morbidity and mortality worldwide as new variant strains evolve and infect susceptible individuals. Development of annual updates to vaccine boosters incorporating new variants is likely (similar to new formulations of annual flu vaccines developed to immunize against the current and predominant circulating strain), requiring testing of protective immunity provided by the updated vaccines. While the main cause of morbidity and mortality associated with SAR-CoV-2 infection is respiratory in nature it has been shown that infection with SARS-CoV-2 virus also affects other organ systems (1-4). Further, it has been shown to directly infect other non-respiratory organs and tissues (5, 6), potentially contributing to non-respiratory related morbidity. In our previous publication (7), we demonstrated that SARS-CoV-2 spike proteins could specifically bind to hepatocytes and immortalized hepatocyte-like cells (HLC) via the asialoglycoprotein receptor-1 (ASGR-1) providing a potential portal for attachment, internalization, and infection of hepatocytes. Hepatocytes/HLC do not express detectable ACE-2, precluding that from being a portal for viral entry. We also demonstrated that the S1 portion of the spike protein was unable to bind to hepatocytes/HLC, barring that from being the mechanism for spike binding and strongly suggesting that S2 was the portion of the spike protein responsible for that binding.

The expression of Transmembrane Serine Protease-2 (TMPRSS-2) on hepatocytes could provide a potential co-factor for internalization of the virus, as it cleaves the S1 and S2 portions of the spike protein and is essential to viral internalization via the ACE-2 portal pathway (8). Inhibition of spike protein binding to hepatocytes/HLC by preincubation of spike proteins with commercially available spike protein-specific monoclonal antibodies suggested that hepatocytes/HLC could provide a useful platform for assessing the effectiveness of antibodies to interfere with S2 binding to cell surface receptors, including ASGR-1. To our knowledge, the development of anti-S2 directed antibodies post-vaccination has not been studied or reported.

To document that immunization with the currently available vaccines elicited an antibody response against the S2-mediated binding to HLC, we collected serum from 10 individuals pre-and post-vaccination and tested them for the development of serum antibodies reactive to variant spike proteins and their capacity to block the binding of spike proteins to HLC. As a positive control, E12-AT-2 cells was used as an additional target cell to follow the concurrent development of serum antibody expression against the already well-documented S1 receptor binding domain (RBD) to ACE-2. Variant-specific antibodies were detected and quantitated by a flow cytometry-based immunofluorescent assay utilizing spike variant proteins bound to paramagnetic beads. The capacity of serum antibodies to inhibit binding of spike proteins to target cells was evaluated by fluorescent confocal microscopy utilizing biotinylated variant spike proteins, pre-incubation of spike proteins with post-vaccination serum, E12-HLC (HLC) and E12-AT-2 (AT-2) target cells. This study suggests that HLC cells can provide a valuable and reproducible tool to evaluate the development of S2-directed immunity elicited by immunization with current and future vaccines.

## Methods and Materials

### Hepatocyte-like Cells (HLC)

Human hepatocyte-like cells (HLC) were developed from the chemical fusion of immortalized E12-MLPC and primary hepatocytes to create a cell with the phenotypic and biological characteristics of fully mature hepatocytes that were immortalized but not transformed (9). These cells, like primary hepatocytes, were also demonstrated to be a target cell for the binding of SARS-CoV-2 spike proteins via binding to ASGR-1 and that this binding could be prevented by commercially available monoclonal antibodies (7). Cells were maintained and expanded using a specific expansion medium composed of Williams Medium E supplemented with 2% fatty acid-free BSA, 1% ITS solution, 5mM hydrocortisone 21-hemisuccinate, FGF basic (20 ng/ml), FGF-4 (20 ng/ml), HFG (40 ng/ml), SCF (40 ng/ml), Oncostatin M (20 ng/ml), BMP-4 (20 ng/ml), EGF (40 ng/ml) and IL-1β (20 ng/ml). Williams Medium E supplemented with 2% fatty acid-free BSA, 1% ITS solution, 5mM hydrocortisone 21-hemisuccinate, FGF basic (20 ng/ml), FGF-4 (20 ng/ml), HFG (40 ng/ml), SCF (40 ng/ml), Oncostatin M (20 ng/ml), BMP-4 (20 ng/ml), EGF (40 ng/ml) and IL-1β (20 ng/ml) (9).

### Alveolar Type-2-Like Cells (AT-2)

Development of AT-2 like cells from MLPC and their utility in the analysis of SARS-CoV-2 spike binding to respiratory cells has been previously reported (10). AT-2 cells developed by this technique produced cells that were phenotypically and biologically identical to AT-2 cells (marketed as Small Airway Epithelial Cells). Immortalized E12-MLPC were differentiated to AT-2-like cells by the same method previously developed for non-immortalized MLPC (11). Briefly, E12-MPLC were cultured in complete SAGM (Lonza, Walkerville, MD, cat# 3118) for 8-10 days to facilitate differentiation to AT-2-like cells, which were then expanded in the same medium for the studies. Cells that were generated (MLPC, HLC and AT-2) and other tissues (cord blood and peripheral blood samples) used in this study received IRB approval by the Quorum Review Protocol #800, March 3, 2005. Donations were collected with donor consent for research use only.

### Spike Proteins

Biotinylated variant spike proteins were used in the development of both the flow cytometry-based quantitative assay and the confocal spike binding/blocking assay. Biotinylated spike variants that were available for the study were the original isolate (RBD SPD-C822E9, ACROBiosystems) and its spike 1 region (S1N-C82E8, ACROBiosystems), the alpha (α) variant (B.1.1.7)(SPNC82E5, ACROBiosystems, Newark, DE), the beta variant (β) (B.1.351)(SPN-C82E6, ACROBiosystems), a combination of the alpha, beta and gamma (γ) variants (B.1.1.7/B.1.351/P1) (SPN-C82E3, ACROBiosystems) and the delta (δ) variant (B.1.617.2)(SPD-C82Ed, ACROBiosystems).

### Quantitative Flow Cytometry Assay Spike Protein Labeled Beads

The detection of spike protein specific antibodies by flow cytometry was enabled by using 7 mm diameter avidin-coated paramagnetic beads (VM-60-10, Spherotech, Forest Lake, IL) with the various biotinylated bound spike proteins. Briefly, saturating concentrations of biotinylated spike proteins (8.3 μg) were reacted with 200 ml of (1% w/v) washed beads. After 2 hours of gently mixing, the beads were washed and resuspended with 6 ml of PBS with 1% BSA for use in the assay. Final concentration of beads was approximately 5 × 10^6^ beads per ml.

### Quantitative Flow Cytometry Assay

The standard curve for quantitation of spike-reactive antibodies was developed utilizing the spike-protein coated beads. The assay procedure was an initial incubation of 100 ml of bead suspension with 100 ml of known concentrations of monoclonal antibody or serum samples for one hour with mixing. Beads were washed and incubated with goat-anti-human IgG-alexa-488 (A48276, Life Technologies) for 30 minutes. Beads were again washed and resuspended for analysis on a Becton-Dickenson FACScaliber flow cytometer on the FL-1 channel. The standard curve was developed for all the variants using the anti-RBD-specific monoclonal antibody developed by ACROBioSystems (SAD-S35). All standard curves were developed using this antibody and the secondary antibody. Mean channel fluorescence of each serum sample was compared to the standard curve to determine the concentration of spike protein-reactive antibodies. While monoclonal antibodies do not reflect on the diverse polyclonal reactivity that would be seen in an immune serum, it does provide a reasonable quantification of total human antibody bound to the spike protein.

### Confocal Immunofluorescent Analysis

The procedure previously used to determine the binding and blocking of spike proteins to HLC and AT-2 targets cells (7, 10) was utilized to determine the ability of post-vaccination serum to inhibit the binding of variant spike proteins. Target cells (200 ml at 1 × 10^6^/ml) were plated in 16 well collagen-coated chamber slides. Cells were allowed to bind to the slides overnight. After overnight incubation, medium was removed, and cells were fixed in 1% formaldehyde for one hour. Cells were then washed twice with 200 ml PrepaCyte permeabilization medium (WBP-1000, CMDG, St. Paul, MN). To determine the binding of each variant spike protein to target cells, cells in each well were incubated with 250 ng of spike proteins diluted in 100 ml of PermaCyte Medium for 30 minutes. Cells were then washed twice with PermaCyte Medium and then incubated with 100 ng of streptavidin-alexa 594 (S11227, Life Technologies, Eugene, OR) and counterstained with DAPI for 30 minutes. Cells were then washed with 200 ml of PermaCyte Medium and analyzed by confocal microscopy. All confocal images were obtained using an Olympus Fluoview 1000 confocal microscope.

To determine the degree of blocking of spike binding to target cells mediated by serum antibodies, we used a modification of the same procedure used to examine binding of the spike protein to the target cells. Briefly, 250 ng aliquots of variant spike proteins were incubated with 100 ml of subject serum for 1 hour with mixing prior to addition to fixed and permeabilized cells. Cells were then incubated for 30 minutes, washed with PermaCyte and then incubated for another 30 minutes with streptavidin-alexa 594 and DAPI. Cells were then washed with 200 ml of Permacyte Medium. Cells were then analyzed by confocal microscopy.

### Cellular Flow Cytometry

The utility of analyzing the binding of biotinylated spike proteins by flow cytometry was examined by using a similar procedure used for the analysis of spike protein binding to slide-plated cells but adapted to cells in suspension. Briefly, cells were directly subjected to similar conditions to those bound to slides (dissociation with expansion flasks by Tryp-LE, washing and resuspension). Cells (5 × 10^5^) were fixed for one hour in 1% formaldehyde; and were then washed and permeabilized with 2 ml of PermaCyte Medium. They were then incubated for 30 minutes with 250 ng of biotinylated spike variants, washed twice with 2 ml of PermaCyte Medium and incubated with streptavidin-Alexa 594 and incubated for 30 minutes. Cells were again washed with 2 ml of Permacyte Medium and resuspended in PBS and analyzed by flow cytometry.

## Results

### Development of Quantitative Flow Cytometry for Anti-Spike Antibodies

As described in the Methods and Materials section above, we utilized the biotinylated spike protein variant coated beads to develop a standard curve utilizing the ACROBiosystems antibody as a standard. While it is important to understand that comparing a monoclonal antibody to a polyclonal antibody response does not account for differences in the binding affinities of individual antibodies in a polyclonal vaccinated serum, nor their binding to different portions of the spike protein, it does provide a good estimate of antibody binding to the variant spike protein at a given concentration. Figure 1 shows a typical standard curve utilizing the ACROBiosytems antibody against spike proteins. The figure also shows the gating on beads and the fluorescent signals on FL-1 that correlate to antibody concentration. This assay, as shown, was used to analyze donor serum to determine total anti-SARS-CoV-2. The sensitivity of this assay was 0.153-2500 ng/ml.

**Figure 1.**
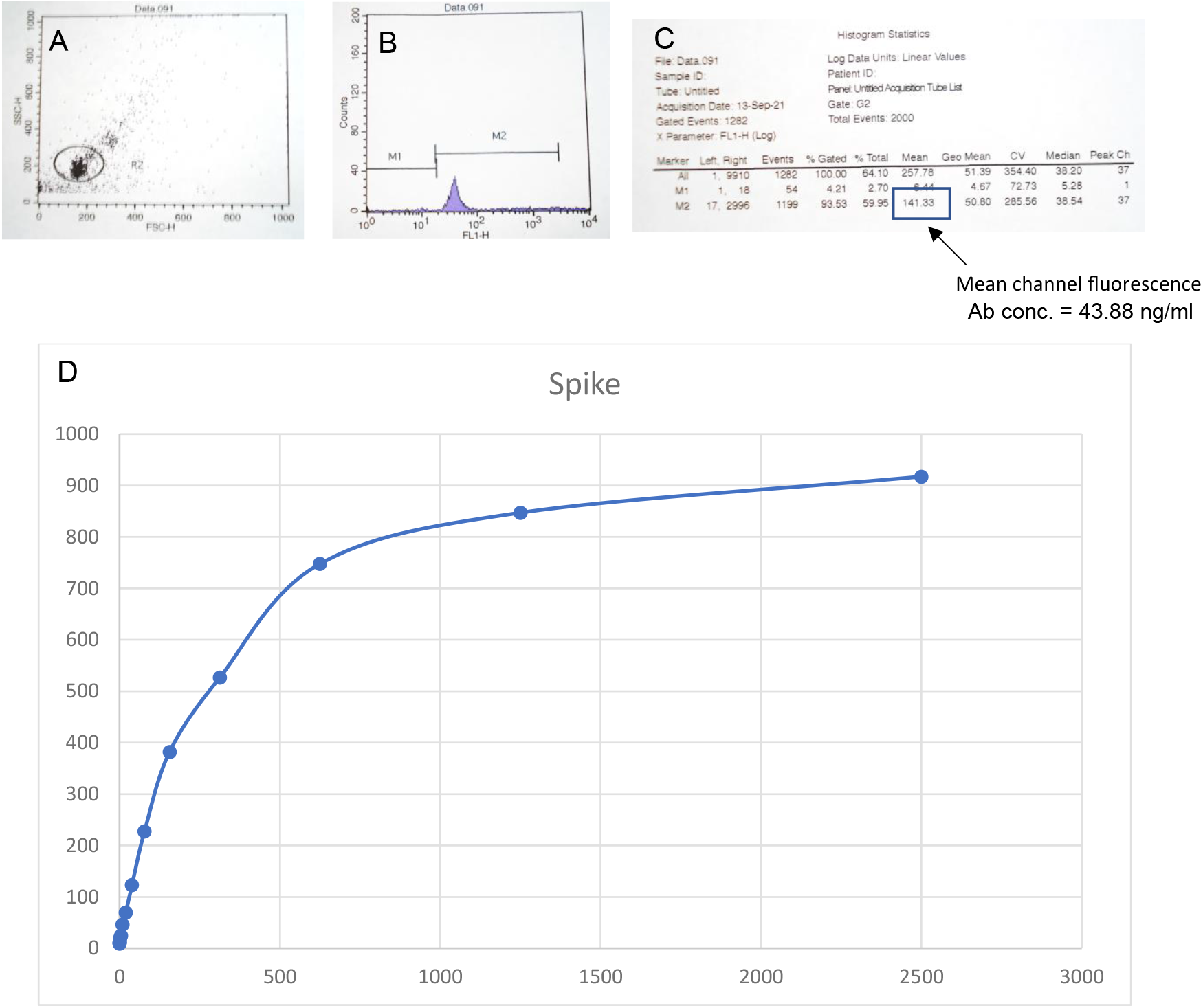
Development of Quantitative Flow Cytometry for Anti-Spike Antibodies. Main population of beads were gated by Forward and Side light scatter (**A**). Binding of antibody to spike protein coated beads was detected by binding of goat-anti-human alexa-488 antibody (**B**). Fluorescent signal correlates to anti-Spike protein-specific antibody concentration (**C**). Standard curves were developed by incubation of beads with known concentrations of neutralizing monoclonal antibody (**D**).

### Comparison of anti-SARS-CoV-2 Antibody Expression Stimulated by the J&J, Moderna and Pfizer Vaccines

As determined by the quantitation flow cytometry assay, levels of spike variant-specific antibodies stimulated by the 3 vaccines were compared at 2.5 months post-vaccination. As shown in Table 1, the highest antibody levels were seen with the Moderna vaccine, while similar levels of antibody were stimulated by the J&J and Pfizer vaccines. In individual samples there was a large variability seen between individuals, and within each there was often a large variability seen between variants. This is shown in the table by the individual results and the large standard deviations.

**Table I.** Quantitation of anti-SARS-CoV-2 serum antibody expressed 2.5 months after vaccination. Data is reported as mean antibody concentration ± standard deviation and individual concentrations for each subject in this study using the quantitative flow cytometry IFA.

| Vaccine | Spike | Spike 1 | Alpha | Beta | Alpha/Beta/Delta | Delta |
| --- | --- | --- | --- | --- | --- | --- |
| J&J | 74.9±29.7 | 65.5±35.4 | 767.3±115.4 | 737.5±454.9 | 230.9±95.9 | 90.3±68.9 |
| Subject 1 | 57.78 | 72.38 | 120.68 | 682.18 | 911.68 | 65.17 |
| Subject 2 | 109.28 | 27.28 | 295.28 | 721.18 | 221.28 | 37.47 |
| Subject 3 | 57.78 | 97.08 | 276.88 | 898.64 | 1079.58 | 168.27 |
| Moderna | 124.2±81.9 | 269.5±164 | 1031.4±1272 | 1237.5±829 | 1365±324.9 | 900.9±1387.9 |
| Subject 4 | 211.08 | 365.58 | 316.08 | 1451.28 | 463.58 | 193.07 |
| Subject 5 | 48.48 | 80.18 | 278.08 | 1006.38 | 748.98 | 9.77 |
| Subject 6 | 113.08 | 362.78 | 2500.08 | 1638.18 | 2500 | 2500 |
| Pfizer | 36.6±15.5 | 78.9±27.1 | 828.9±309.5 | 853.9±703.9 | 190±207.5 | 136.3±248.7 |
| Subject 7 | 31.88 | 93.18 | 30.48 | 707.98 | 815.08 | 509.37 |
| Subject 8 | 43.88 | 51.88 | 204.28 | 533.28 | 670.08 | 9.4 |
| Subject 9 | 53.08 | 60.78 | 46.98 | 815.08 | 120.68 | 9.77 |
| Subject 10 | 17.38 | 109.58 | 478.28 | 1259.18 | 1809.68 | 16.77 |

### Binding of Spike Proteins to HLC and AT-2

The binding of SARS-CoV-2 spike proteins to HLC and AT-2 was determined by fluorescent confocal microscopy. Positive binding of spike protein was demonstrated by red fluorescence. As shown in Figure 2, HLC did not bind spike 1, but was able to bind the wild type and all the variant spike proteins. AT-2 cells were able to bind each of the different spike proteins.

**Figure 2.**
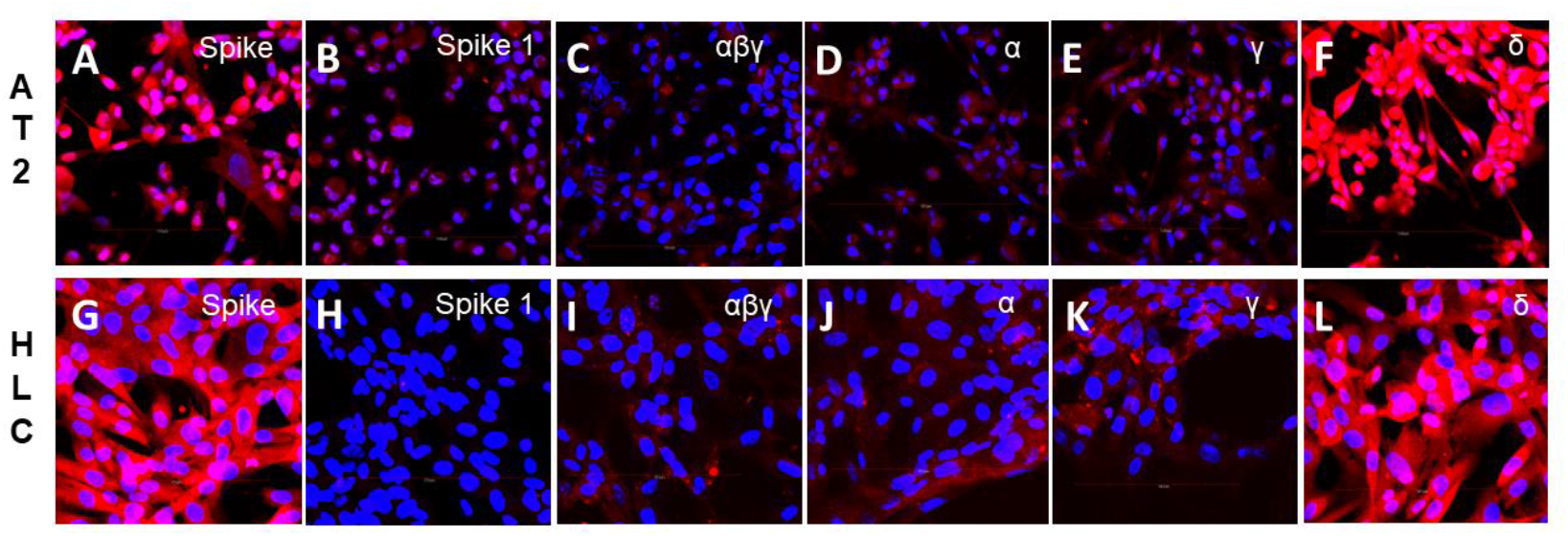
Binding of Wild Type and Variant Spike Proteins to HLC and AT-2-like Cells. Binding of Spike proteins to AT-2 like cells is shown in Figures **A**-**F**. Binding of Spike proteins to HLC is shown in Figures **G**-**L**. Binding of Wild Type Spike protein is shown in Figures **A** and **G**. Binding of Spike 1 protein is shown in Figures **B** and **H**. Binding of α/β/γ variant is shown in Figures **C** and **I**. Binding of α variant is shown in Figures **D** and **J**. Binding of γ variant is shown in Figures **E** and **K**. Binding of δ variant is shown in Figures **F**and **L**. Binding of Spike proteins to the cells is demonstrated by red fluorescence. As shown, Spike 1 protein does not bind to HLC.

### Analysis of Spike-Blocking Antibodies

The presence of antibodies capable of inhibiting binding of spike proteins was determined by fluorescent confocal analysis. Spike proteins were pre-incubated with subject serum prior to addition to target cells. As was seen in Figure 2, positive binding of Spike proteins was indicated by red fluorescence of cells. Inhibition of binding was shown by reduced or absent red fluorescence of the cells.

Blocking antibody against the spike variants developed from 2 weeks to 2.5 months as shown with both target HLC (Figure 3) and AT-2 cells (Figure 4). There was significant, but not complete block of spike protein binding by serum at 2 weeks, while at 2.5 months there was complete block of spike binding.

**Figure 3.**
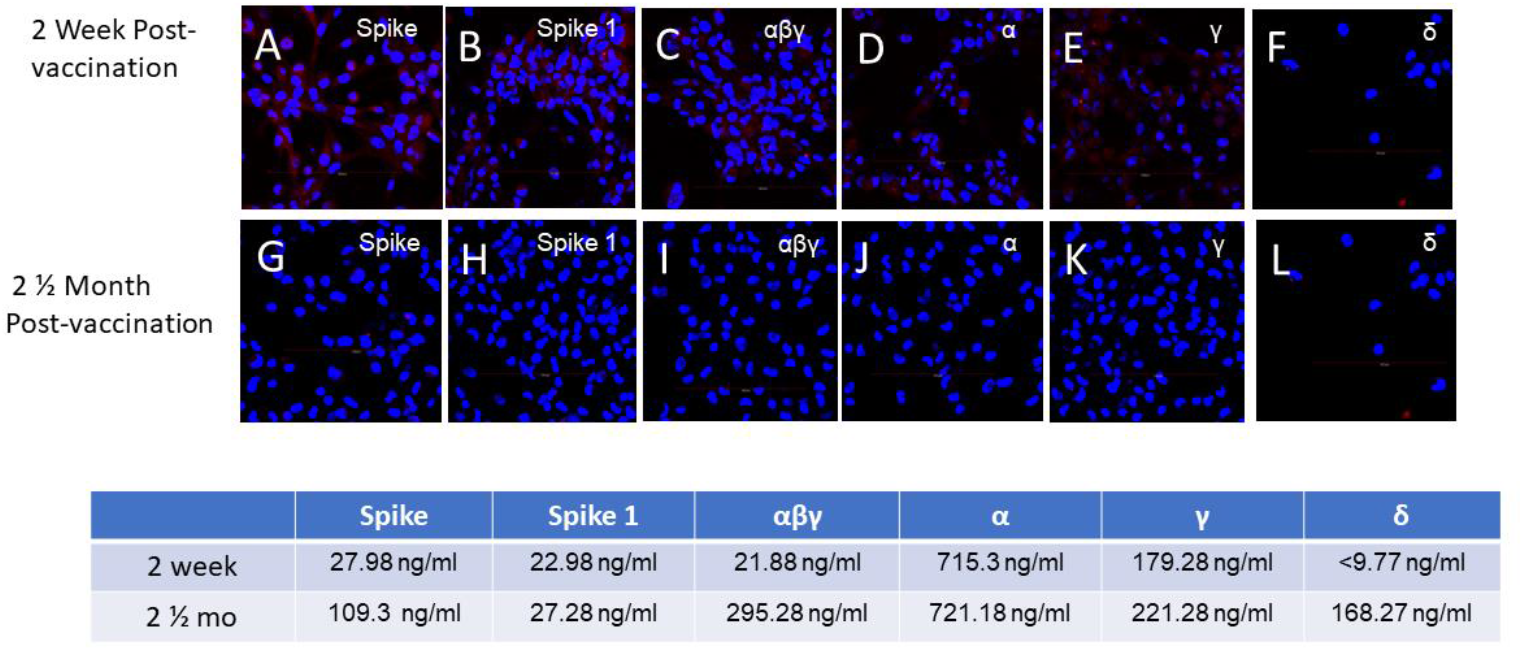
Development of Spike-blocking Antibody Post-Vaccination. Spike proteins were incubated with post-vaccination serum prior to addition to HLC. Expression of red fluorescence demonstrates binding of spike protein to cells. Lack of red fluorescence demonstrates complete block of spike binding. Block of spike protein binding by serum 2 weeks post-vaccination is shown in Figures **A**-**F**. Block of spike protein binding by serum 2.5 months post vaccination is shown in Figures **G**-**L**. Spike-protein specific antibody concentrations determined by quantitative flow cytometry IFA at both 2 weeks and 2.5 months are shown below. Antibodies concentrations were lower in the 2 week serum than the 2.5 month serum and were less able to completely block the binding of the spike proteins. The results shown were for the J&J vaccine, but similar results were seen with the Moderna and Pfizer vaccines.

**Figure 4.**
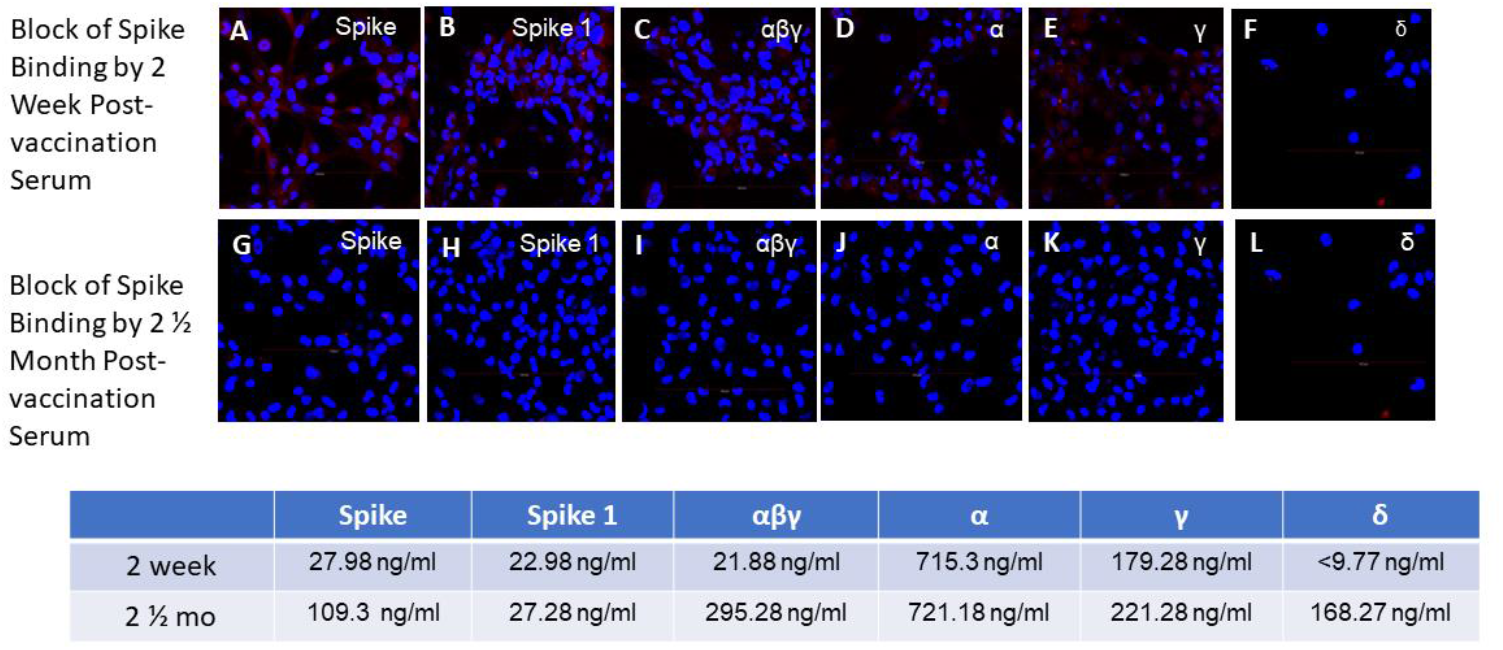
Development of Spike-blocking Antibody Post-Vaccination. Spike proteins were incubated with post-vaccination serum prior to addition to AT-2 cells. Expression of red fluorescence demonstrates binding of spike protein to cells. Lack of red fluorescence demonstrates complete block of spike binding. Block of spike protein binding by serum 2 weeks post-vaccination is shown in Figures **A**-**F**. Block of spike protein binding by serum 2 ½ months post vaccination is shown in Figures **G**-**L**. Spike-protein specific antibody concentrations determined by quantitative flow cytometry IFA at both 2 weeks and 2 ½ months are shown below. Antibodies concentrations were lower in the 2 week serum than the 2 ½ month serum and were less able to completely block the binding of the spike proteins. The results shown were for the J&J vaccine, but similar results were seen with the Moderna and Pfizer vaccines.

Analysis of variant-specific blocking antibodies was analyzed for serum obtained from individuals receiving the different vaccinations. As seen in Figure 5, pre-vaccination serum did not block spike protein binding to HLC, whereas each of the different vaccinations stimulated the expression of blocking antibodies to all the variants. AT-2 cells were able to bind each of the different spike proteins. Binding to AT-2 cells were also uniformly blocked by serum from individuals receiving any of the 3 vaccines (Figure 6). Adaptation of this assay for flow cytometry demonstrated that this system worked well for determining spike binding to target cells. An additional desirable quality afforded by flow cytometry is the ability to easily compare the relative fluorescence of each binding which directly relates to the amount of spike protein that is bound. (Figure 7). These cellular targets were demonstrated to be useful in these analyses.

**Figure 5.**
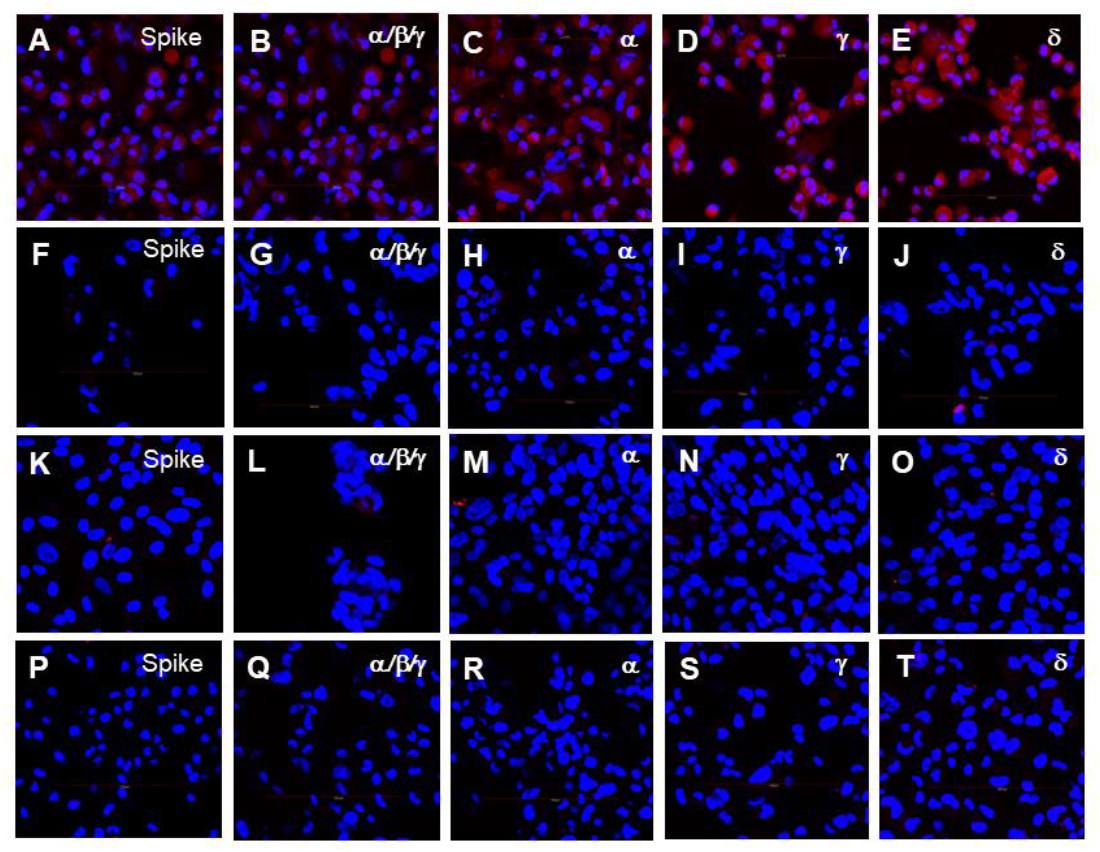
Vaccination-Specific Expression of Spike-blocking Antibody at 2.5 months post-vaccination. Spike proteins were incubated with spike proteins prior to addition to HLC. Block of spike proteins with prevaccination serum is shown in Figures **A**-**E**. Block of spike proteins with post-Moderna vaccination serum is shown in Figures **F**-**J**. Block of spike proteins with post-J&J vaccination serum is shown in Figures **K**-**O**. Block of spike proteins with post-Pfizer vaccination serum is shown in Figures **P**-**T**. As shown, all vaccinations were capable of completely blocking spike protein binding.

**Figure 6.**
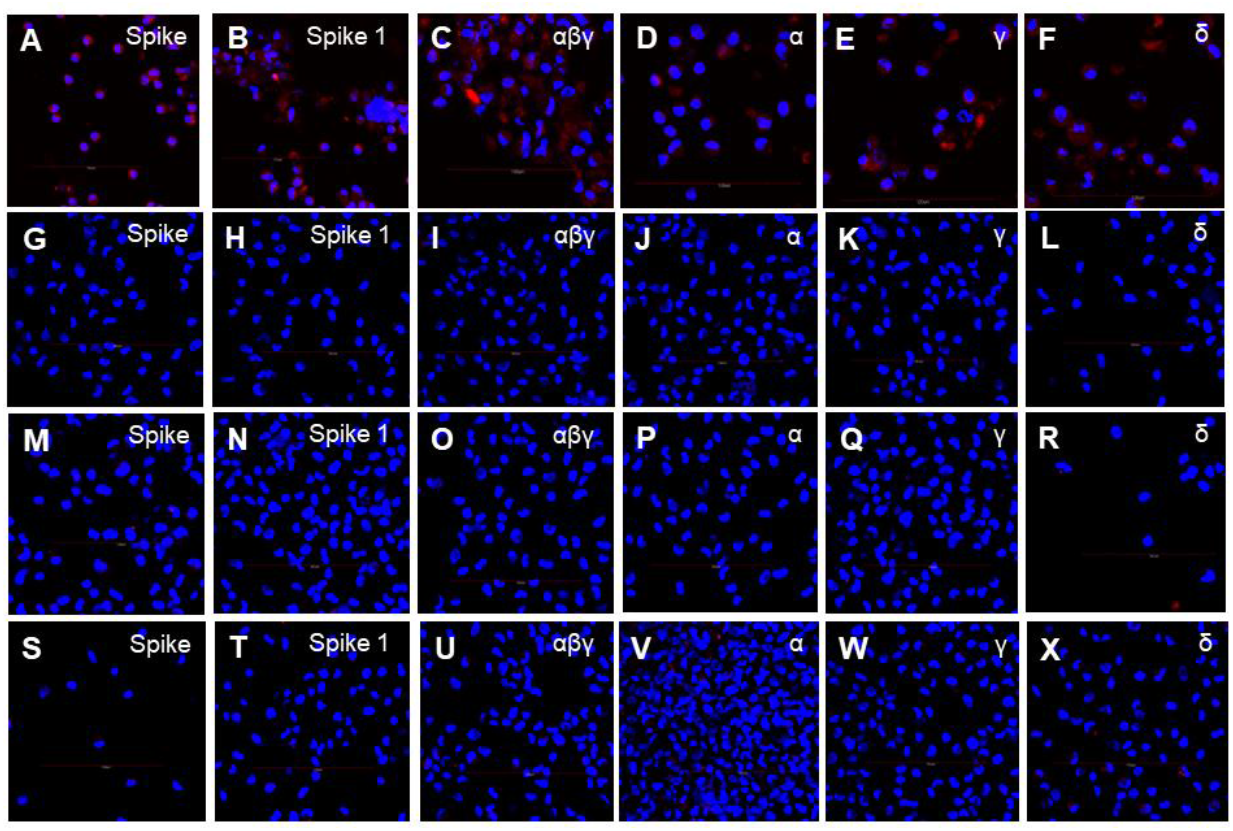
Vaccination-Specific Expression of Spike-blocking Antibody at 2.5 months post-vaccination. Spike proteins were incubated with spike proteins prior to addition to AT-2. Block of spike proteins with pre-vaccination serum is shown in Figures **A**-**F**. Block of spike proteins with post-Moderna vaccination serum is shown in Figures **G**-**L**. Block of spike proteins with post-J&J vaccination serum is shown in Figures **M**-**R**. Block of spike proteins with post-Pfizer vaccination serum is shown in Figures **S**-**X**.

**Figure 7.**
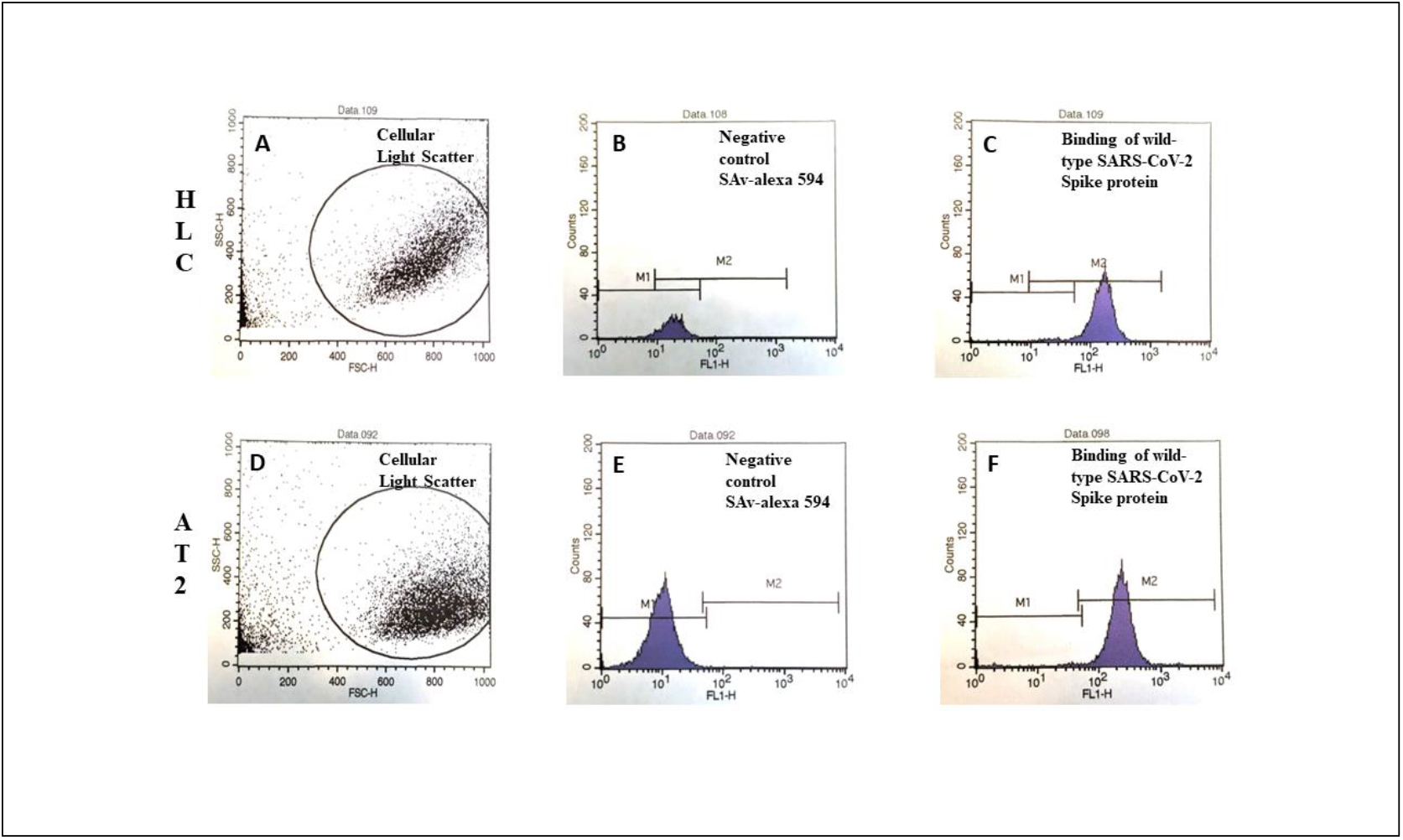
Flow Cytometric Analysis of Spike Binding to HLC and AT-2. Gating on HLC based on Forward and Side light scatter is shown in Figure **A**. Gating on AT-2 based on Forward and Side light scatter is shown in Figure **D**. Background fluorescence was determined by non-specific binding of Alexa-594, Figures **B** and **E**. Binding of the Wild Type Spike proteins is shown in Figures **C** and **F**.

## Discussion

The SARS-CoV-2 virus is spread primarily by airborne transmission. The respiratory system is the first entry point for the virus, via the ACE-2 receptor, which is the attachment point for the virus (12, 13). The respiratory system is the main focal point for most of the morbidity and mortality associated with infection (14). In some cases, infection with the virus has also been associated with non-respiratory pathology including brain, kidney, cardiac, digestive system, and liver (1-6). While many organ systems contain cells that express ACE-2 and can be infected by that portal, it has been demonstrated that the SARS-CoV-2 virus is capable of infecting cells that do not possess the ACE-2 receptor (5, 7). It has been shown that hepatic parenchymal cells could be infected by the virus, yet hepatocytes do not express ACE-2 (7, 8), suggesting a different mechanism for viral attachment and entry.

In a previous publication (7), we reported that hepatocytes do not express ACE-2 but still bound the viral spike protein. We also reported that hepatocytes were unable to bind the S1 portion, which contains the receptor binding domain (RBD) that binds to ACE-2. Similarly, we showed that an immortalized HLC also did not express ACE-2, yet bound spike protein and not the S1 protein. Additionally, we demonstrated that antibodies directed against the surface membrane asialoglycoprotein receptor could block the binding of spike protein to hepatocytes and HLC. Combined, these results suggested that the spike protein binds to hepatocytes and HLC via the asialoglycoprotein-1 receptor and the S2 portion of the spike protein. The additional presence of TMPRSS-2 on hepatocytes (8) likely provides a co-factor needed for internalization of the virus. The block of spike protein binding by commercially available anti-spike monoclonal antibodies to hepatocytes and HLC suggests that HLC could serve as a target cell to determine the ability of post-vaccination serum to block the binding of spike protein, thus serving as a useful tool to assess immunity against the S2 portion of the spike protein. We have previously reported differentiation of alveolar type-2 cells (AT-2) from immortalized E12-MLPC cells as an *in vitro* system to study the binding of spike protein to the ACE-2 receptor (10). The model provides a cell line that can assess immunity to the S1 portion of the spike protein. The use of two different cell types from lung and liver enables the ability to assess antibodies directed towards both the S1 (extensively studied) and the S2 (rarely studied) portions of the spike protein.

This paper describes a study utilizing the HLC and AT-2 as target cells to determine if binding to the spike protein could be inhibited by post-vaccination serum. The serum of ten individuals were obtained prior to and post vaccination with the three different vaccines (J&J, Moderna, Pfizer). Serum samples were quantitated for anti-spike specific antibody by flow cytometry-based IFA (Figure 1 and Table 1), and tested for their ability to inhibit the binding of spike protein to the target cells. Samples were also tested for the ability to block spike protein binding using the same methodology previously described to study both the binding and blocking of spike proteins utilizing HLC and AT-2 cells (7, 10). We observed that all variants tested bound to both HLC and AT-2 cells (we had one opportunity to test the omicron variant and found that it bound to both HLC and AT-2 cells and could be blocked by immune serum, data not shown). We also established that serum from all subjects tested after vaccination with all three vaccines stimulated the production of antibodies that completely blocked the binding of all the spike variants. When the antibody response was analyzed, we identified significant variations in antibody levels between individuals in addition to within each individual themselves. Antibody levels did not necessarily reflect the degree of spike protein binding block to target cells, as even low levels of antibody were capable of complete inhibition of binding. We believe that the spike-blocking assay provided a better assessment of protective immunity than just measuring antibody levels. While not extensively studied, we found that the binding/inhibition assays were easily adaptable to analysis by flow cytometry allowing utilization of this assay to laboratories that don’t have access to confocal microscopy but do have availability of a flow cytometer.

We believe that this assay has many advantages over simply determining gross anti-SARS-CoV-2 antibody levels or the use of pseudo-viruses to test inhibition of binding. First was the development of HLC and AT-2 cell lines that are stable, immortalized, highly differentiated and express the phenotypic and biological properties of mature and fully functional primary human hepatocytes and primary small alveolar cells, respectively. The use of biotinylated spike proteins to study binding of the virus to target cells instead of live virus or pseudo-virus also provides a significant advantage in testing. The use of this combined system utilizes immortalized cell lines that can be readily grown and maintained to provide a stable and reproducible source of target cells. The use of commercially available spike protein also eliminates the need for creation of new pseudo-viruses with the evolution of each new variant or use of the actual live virus. This eliminates any dangers involved with working with live or pseudo-viruses and the need for extensive biological containment. These features allow this assay to be produced in large-scale and safely utilized by any normally equipped analytical laboratory. We believe that our system should be widely utilized as it provides a more accurate assessment of actual protective immunity than just quantitation of SARS-CoV-2-specific antibodies and is safe and easily implementable in standard analytical laboratories.

## Disclosure

This study was funded in its entirety by CMDG LLC. The authors report no other conflicts of interest in this work.

## Data Availability

All data produced in the present work are contained in the manuscript

